# Functional analysis enables reclassification of *SCN1A* variants of uncertain significance in epilepsy

**DOI:** 10.64898/2026.11.07.26367252

**Authors:** Keerthi Pilli, Talia J. Allan, Mark F. Bennett, Rebekah V. Harris, Amy L. Schneider, Qianyi Shen, Joanne G. Ma, Michael S. Hildebrand, Elizabeth E. Palmer, Alfred L. George, Samuel F. Berkovic, Jamie I. Vandenberg, Ingrid E. Scheffer, Chai-Ann Ng

**Author notes:** These authors contributed equally to this work (Jamie I. Vandenberg, Ingrid E. Scheffer, Chai-Ann Ng). Correspondence to: Chai-Ann Ng Victor Chang Cardiac Research Institute, 405 Liverpool St, Darlinghurst, NSW 2010, Australia. Correspondence may also be addressed to: Ingrid Scheffer Epilepsy Research Centre, 245 Burgundy Street, Heidelberg, Victoria 3084, Australia. Jamie Vandenberg Victor Chang Cardiac Research Institute, 405 Liverpool St, Darlinghurst, NSW 2010, Australia.

## Abstract

*SCN1A,* the most prominent gene for epilepsy, encodes the pore-forming subunit of the voltage-gated sodium channel, Na_V_1.1. Loss-of-function variants reduce Na_V_1.1 activity in inhibitory neurons leading to hyperexcitability; they are associated with a spectrum of epilepsies, from self-limited phenotypes in genetic epilepsy with febrile seizures plus, to the prototypic developmental and epileptic encephalopathy, Dravet syndrome. Thousands of *SCN1A* missense variants have been identified, but determining pathogenicity using ACMG criteria can be challenging. Currently, 48% of missense *SCN1A* variants in ClinVar are classified as variants of uncertain significance (VUS).

We aimed to provide functional evidence to inform variant classification using a high-throughput automated patch-clamp assay. We validated our assay using 27 true pathogenic *SCN1A* variants and 25 true benign variants. The results show that 17/27 true pathogenic variants from patients with Dravet syndrome showed severe reductions in current density, precluding gating analysis. Of the remaining ten true pathogenic variants, seven showed an increased voltage difference between the midpoints of activation and inactivation (*V*_0.5diff_). Current density and *V*_0.5diff_ were combined within a graded Bayesian framework to derive a functional log-likelihood ratio reflecting the magnitude of loss of function.

We then applied our validated assay to 16 *SCN1A* missense variants detected in an Australian cohort: three likely pathogenic, one likely benign and 12 VUSs. These variants occurred in 31 individuals from 16 families with a broad phenotypic spectrum, including four individuals with Dravet syndrome who carried p.Ser243Phe, p.Met973Val or p.Lys1911Glu. All three variants showed loss of function: p.Ser243Phe had reduced current density, whereas p.Met973Val and p.Lys1911Glu had abnormal *V*_0.5diff_ despite current density within the normal range. Of these 16 variants, ten have combined functional scores consistent with abnormal functional evidence.

Combining our functional evidence with ACMG criteria, our findings changed the classification of eight of 16 variants (50%), with six of 12 VUS reclassified as likely pathogenic and two of three likely pathogenic variants upgraded to pathogenic. We show that calibrated automated patch-clamp assays can provide actionable functional data to enable the interpretation and reclassification of *SCN1A* variants and inform clinical care, allowing therapeutic optimisation and facilitating access to genetic therapies.

## Introduction

*SCN1A* encodes the pore-forming subunit of the Na_V_1.1 voltage-gated sodium channel, which is predominantly expressed in inhibitory GABAergic interneurons.^1^ Loss-of-function (LOF) of Na_v_1.1^2^ reduces inhibitory signalling, which leads to hyperexcitability of the central nervous system.^1,3^ Pathogenic LOF variants in *SCN1A* underpin a spectrum of epilepsies, ranging from developmental and epileptic encephalopathies (DEEs) such as Dravet syndrome,^2–5^ to genetic epilepsy with febrile seizures plus (GEFS+).^6–8^ Dravet syndrome, the prototypic DEE, is caused by pathogenic LOF variants in *SCN1A* in more than 90% of patients.^9^ By contrast, GEFS+ is a clinically heterogeneous, autosomal dominant familial epilepsy encompassing a spectrum of epilepsy phenotypes, with febrile seizures as a hallmark; notably, only 19% of GEFS+ families harbour pathogenic *SCN1A* variants.^8^

Variant pathogenicity is determined in accordance with the guidelines established by the American College of Medical Genetics and Genomics (ACMG) and the Association for Molecular Pathology (AMP),^10^ which classify genetic variants as pathogenic, likely pathogenic, variants of uncertain significance (VUS), likely benign or benign. These genetic classification guidelines have facilitated diagnosis across numerous genetic disorders.^11^ For rare diseases, however, it can be challenging to obtain sufficient evidence to reach a definitive variant classification. This issue becomes more pronounced for missense variants, with more than 1.2 million missense variants currently classified as VUS in ClinVar,^12^ including 1,315 of 2,735 missense *SCN1A* variants (accessed 04 August 2026). Accurate reclassification of *SCN1A* VUS has critical clinical utility; sodium channel-blockers should be avoided in individuals with pathogenic LOF *SCN1A* variants. Patients may also become eligible for *SCN1A* precision medicine genetic therapies and clinical trials.

Functional evidence, such as that obtained through patch-clamp electrophysiology studies can be useful for reclassification of VUS in ion channel disorders.^13–17^ The Clinical Genome Resource (ClinGen) Sequence Variant Interpretation (SVI) Working Group recently recommended that functional assays need to be validated against a minimum of 19 true pathogenic variants and 19 true benign variants, to achieve a strong level of evidence for variant classification.^18^ Gene-specific classification specifications for the neuronal sodium channel genes, including SCN1A, have since been issued by the ClinGen Epilepsy Sodium Channel Variant Curation Expert Panel.^19^ The generation of patch-clamp data for such a large number of variants in ion channel genes has become feasible due to the advent of automated patch clamp (APC) technology, which has enabled 100 times the throughput of functional data collection for ion channel variants.^17,20–24^

Here, we determined the sensitivity and specificity of an APC assay for detecting LOF variants in *SCN1A* and evaluated its utility in reclassifying *SCN1A* missense VUS in an Australian cohort. Improving variant classification with robust functional evidence supports a definitive diagnostic framework regarding aetiology, enabling targeted management and better long-term outcomes.

## Materials and methods

### Variant information

Twenty-five true benign *SCN1A* variants were selected using a Grpmax Filtering Allele Frequency (FAF) greater than 0.005% from gnomAD v4.1. The twenty-seven recurrent *SCN1A* missense variants found in patients with Dravet syndrome were curated as true pathogenic variants.^9^ These true benign and true pathogenic variants are listed in Supplementary Table 1. Sixteen *SCN1A* missense variants detected in patients with epilepsy enrolled in the Genetic Basis of Epilepsy Research Program at The University of Melbourne (Austin Health HREC #H2007/02961) were functionally characterised. All variants were classified using ACMG guidelines supplemented with ClinGen recommendations (Supplementary Table 2). Variants were tested in DNA from family members where available.

### Generation of *SCN1A* plasmids and Flp-In T-REx HEK293 cell lines

WT *SCN1A* (NM_001165963.4) sequences^25^ and *SCN1B* (NM_001037.5), separated by an internal ribosome entry site (IRES), were synthesised and subcloned into the pcDNA^TM^5/FRT/TO plasmid (Thermo Fisher Scientific) by GenScript Inc. (Piscataway, USA). This WT *SCN1A*-IRES-*SCN1B* plasmid was used as template to produce all 68 *SCN1A* variants using site-directed mutagenesis service (GenScript Inc). All variant plasmids were confirmed by Sanger sequencing. The 69 plasmids (WT and 68 variants) were each co-transfected with the Flp recombinase plasmid pOG44 into Flp-In T-REx HEK293 parental cells. Flp-mediated recombination directs the construct to a single FRT site at the same genomic locus in every line, so the resulting hygromycin-resistant populations are isogenic and no single-colony selection is required. Stable lines were selected with 200 μg/mL hygromycin B (Sigma-Aldrich) as described in detail previously.^26^

### APC assay

*SCN1A* expression was induced with 200 ng/mL doxycycline (Sigma-Aldrich) 48 hours before cells were harvested for APC experiments, using Accumax (Sigma-Aldrich, cat. # A7089). Whole-cell patch clamp experiments were performed in 384-well plates using the SyncroPatch 384PE (Nanion Technologies, Munich, Germany) APC system. A single voltage protocol was designed to enable measurement of current density and voltage-dependence of activation and inactivation. For each sweep, the voltage was held at –120 mV for 200 ms, then stepped up to voltages in the range –100 to +60 mV, increasing in 5 mV intervals, for 100 ms, then stepped up to zero mV for 50 ms. The current density was taken from the peak current measured at zero mV after holding at –70 mV to mimic the resting membrane potential of neurons. Each plate contained 32 wells for WT control, 32 wells for each of the ten variants and 32 wells for negative control cells. Each experiment was repeated once on the same day and two plates were run again on a different day using a different batch of cells to ensure the results were reliable and reproducible.

### Data analysis

Data from the APC experiments were analysed using DataControl384 and a customised MatLab script. Initial quality control (QC) parameters implemented in the DataControl384 software were: minimum seal resistance of 500 MΩ, capacitance value between 2–30 pF, series resistance less than 20 MΩ. For gating analysis, secondary QC parameters applied were: time to peak at zero mV between 0.5–1.1 ms for current density measurements, and current amplitude cut-offs of 400 pA to 4 nA for voltage-dependence of activation and inactivation to restrict fitting to well-resolved currents while avoiding series resistance voltage error.

Current density was calculated by dividing the inward sodium current of a cell by its capacitance. The variants were normalised to the mean of the WT of their respective plate to account for variability across plates. As raw current density measurements show a non-Gaussian distribution (see Fig. 1D), a square root transformation was applied to achieve Gaussian distribution of data.

**Figure 1.**
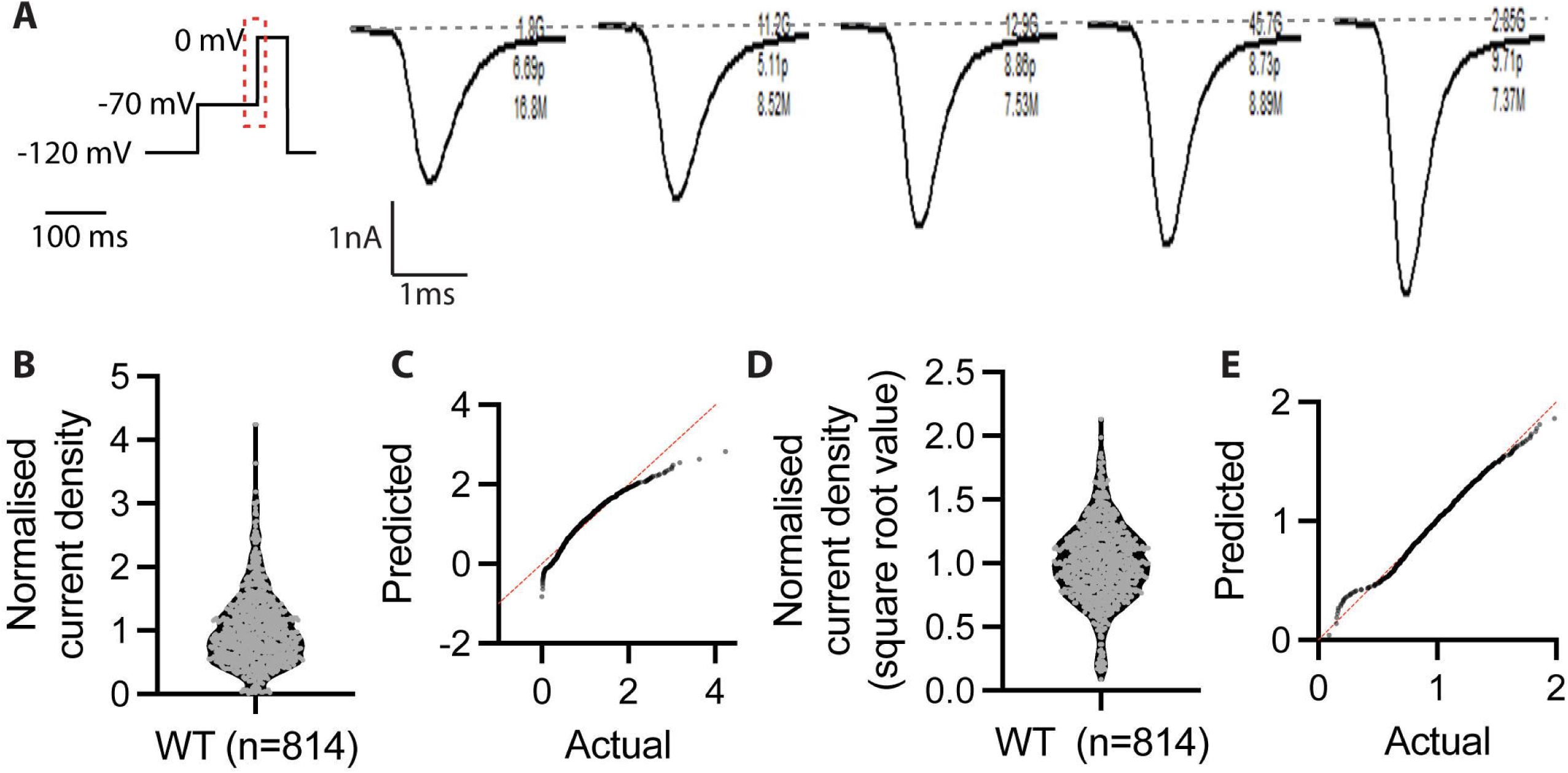
Electrophysiological recording of WT Na_V_1.1 obtained using automated patch clamp platform. (**A**) Representative current traces of WT Na_V_1.1 are highlighted by the red box in the voltage protocol. **(B)** The violin plot for WT cells displaying the distribution of current density measurements is not normally distributed with the corresponding QQ plot **(C). (D)** The violin plot for the same WT cells displays normal distribution after square root transformation with the corresponding QQ plot **(E)**.

Power calculations for sample sizes were performed based on the *SCN1A* cells that passed QCs in the WT-only experiments using the following formula^27^:

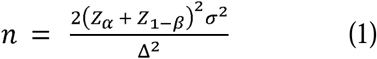

where *n* is the required sample size, *Z*_α_ is a constant for α-error, *Z*_1–β_ is a constant for power of the study, σ is the standard deviation (SD) and Δ is the estimated effect size.

For the voltage-dependence of activation (see Fig. 4A), peak currents were transformed to conductance using the following equation:

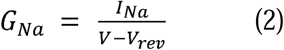

where *G*_Na_ is sodium conductance, *I*_Na_ is the peak sodium current, *V* is the voltage at which peak depolarisation occurs and *V*_rev_ is the reversal potential.

The conductance at each voltage step was normalised to the maximum conductance of the cell (*G/G*_max_) to generate an activation curve. A Boltzmann function was fitted to the conductance data to obtain *V*_0.5a_:

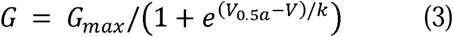

where *G* is conductance, *G*_max_ is the maximum conductance, *V*_0.5a_ is the voltage where half the channels are activated, *V* is the test potential and *k* is the slope factor.

For the voltage-dependence of inactivation (see Fig. 4B), the current at each voltage step was normalised to the maximum current of the cell (*I/I*_max_) to generate an inactivation curve and fitted to a Boltzmann function:

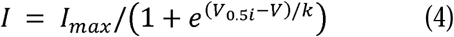

where *I* is current, *I*_max_ is the maximum current, *V*_0.5i_ is half-maximum inactivation voltage, *V* is the test potential and *k* is the slope factor.

We also quantified the difference between the midpoint of channel activation (*V*_0.5a_) and inactivation (*V*_0.5i_) from the same cell:

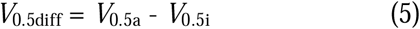

A Z-statistic was employed to establish the functionally normal range for each parameter as the mean ±2 SD of WT and the 25 true benign variants. Current density and *V*_0.5diff_ were combined into a single functional score using Firth-penalised logistic regression on the 25 true benign and 27 true pathogenic variants. A penalty was required because the two classes were completely separated. For the 18 variants in which severe loss of current precluded Boltzmann fitting, *V*_0.5diff_ was set to the WT mean of 25.52 mV before fitting, leaving current density as their only source of evidence. With the intercept omitted because it is absorbed into the calibration step, S = −14.8321 × CD + 1.6296 × *V*_0.5diff_, where CD is the square-root-transformed current density normalised to the plate WT mean, and *V*_0.5diff_ is in mV; a complete-case refit excluding the assigned variants (n = 35) gave S = −16.1594 × CD + 1.6694 × *V*_0.5diff_ and left every calibrated evidence tier unchanged.

The log-likelihood ratio (LLR) framework established for multiplexed assays of variant effect (MAVE),^28^ and applied previously to ion-channel functional assays,^29^ was used for calibration, with an assay-specific modification to the kernel density estimation step. The benign reference set comprised the 25 true benign variants together with WT (n = 26), and the pathogenic reference set comprised the 27 true pathogenic variants. Probability densities of the functional score were estimated separately for each reference set by Gaussian kernel density estimation. Because the two reference distributions do not overlap (benign, S = 23.3– 31.8; pathogenic, S = 34.2–50.0), class-specific bandwidths produce an LLR curve that rises almost discontinuously across the unsampled interval between them, assigning the maximum attainable evidence strength to every reference variant. A single bandwidth was therefore derived by Silverman’s rule of thumb from the pooled benign and pathogenic scores (h = 2.82) and applied to both densities. This represents deliberate smoothing relative to the class-specific values (h = 1.07 for benign and 0.58 for pathogenic) and was adopted to yield a conservative calibration. Thresholds were reported only within the score interval spanned by the reference variants (S = 23.3–50.0), so that no evidence was assigned by extrapolation beyond values supported by the assay.

For each functional score, the likelihood ratio of pathogenicity was calculated as the ratio of the estimated pathogenic density to the estimated benign density and expressed as log_10_ LR. To prevent the ratio from being determined by near-zero density estimates in sparsely sampled regions, both densities were floored at a common minimum equal to 10^-4^ of the density estimated at a pseudo-observation placed ten bandwidths below the lowest reference score, and the resulting profile was constrained to be non-decreasing in the functional score. Evidence thresholds were taken from the Bayesian point system underlying the ACMG/AMP classification framework,^30,31^ as adopted by the ClinGen Sequence Variant Interpretation Working Group, and were labelled with the corresponding functional-evidence codes BS3 and PS3.^10,18^ On the combined functional score these thresholds correspond to S ≥ 34.6 for supporting, S ≥ 35.5 for moderate, S ≥ 37.1 for strong and S ≥ 40.1 for very strong evidence of abnormal function. Although the calibrated curve attains log_10_ LR values consistent with very strong evidence in both directions (thresholds log_10_ LR = ±2.54), pathogenic functional evidence contributed by this assay was capped at strong (PS3_strong) in accordance with ClinGen recommendations for PS3/BS3 assay calibration.^18^ Normal function evidence (BS3) was not applied at any strength (see Limitations).

## Results

### Current density measurement of WT Na_V_1.1 encoded by *SCN1A*

There is considerable cell-to-cell variability in WT Na_V_1.1 current density, as illustrated by the representative current traces measured during a step depolarisation to zero mV from a holding potential of –70 mV (Fig. 1A). The variation observed is similar to that seen for other ion channels, such as Na_V_1.5 and K_V_11.1.^15,16^ The normalised current density of WT at zero mV across six measurement plates (*n*=814) shows a non-normal distribution (Fig. 1B and C). However, the data can be transformed to a normal distribution by using a square-root function. To account for any plate-to-plate variation, current density measurements were normalised to the mean WT from the same plate (Fig. 1D and E). The normalised SD for WT current density was 0.30. Thus, a minimum sample size of 30 was needed to detect a 25% difference in current density, with 90% power and α of 0.05. Alternatively, detecting a 50% difference, with 90% power and α of 0.05, would require 8 cells.

### Normal and abnormal functional range based on true benign and pathogenic *SCN1A* variants

The normalised current densities for the 25 true benign variants (after amalgamating data from all replicate plates) are summarised in Fig. 2A. The violin plot illustrates the distribution of all data for each variant, while the whiskers denote the 5^th^ and 95^th^ percentiles. The mean of the 25 true benign variants was 0.9 ± 0.1, which was used to establish the normal function range, defined by a Z-score between 2 and –2 (Fig. 2B). This allows the normal biological variation to be captured to ensure the difference detected by the assay is not only statistically significant, it is also biologically significant. For the 25 true benign variants, the current density for one of them falls below the –2 threshold (p.Arg1575Cys, Z = – 2.4). Thus the specificity of the assay is 96%. We next measured the current densities of the 27 true pathogenic *SCN1A* variants from patients with Dravet syndrome (see Fig. 3A for examples of current traces from selected pathogenic variants). Twenty-two true pathogenic variants had a mean current density below the normal range established using the 25 true benign variants. This is consistent with current density being the major determinant of the LOF phenotype. There were, however, five true pathogenic variants with current densities that fell within the normal range (Fig. 3B).

**Figure 2.**
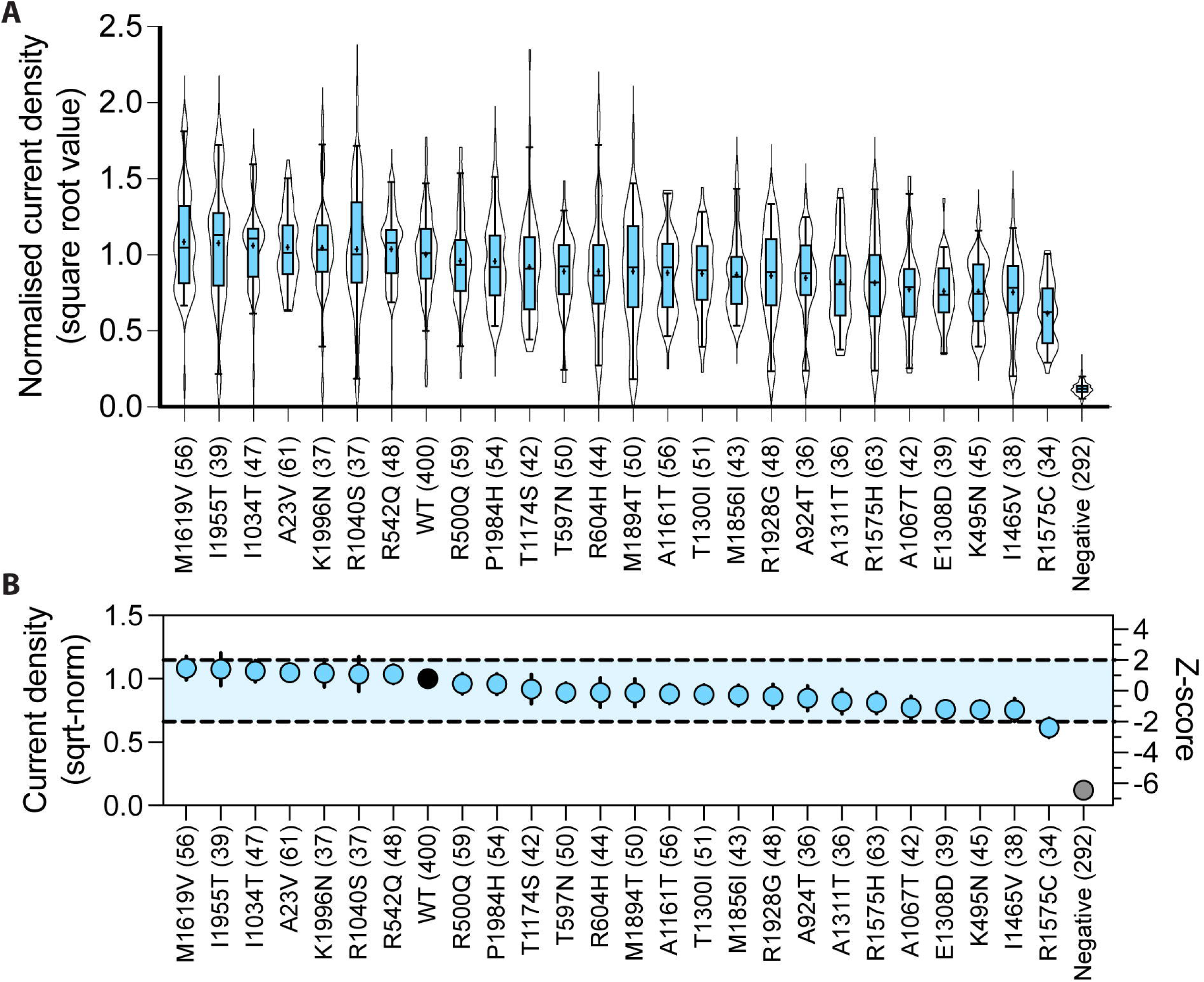
Establishing functionally normal range for *SCN1A* APC assay by current density measurement. **(A)** Summary of current density of 25 true benign variants. All data points that pass all quality control parameters were included. Violin plots show the distribution of all cells; the overlaid box shows the median (line), mean (+) and 5th–95th percentiles (whiskers). **(B)** Normal current density range defined by using Z ±2, corresponds to a square-root, normalised current density of 0.66 - 1.15. Data presented as mean ± 95% CI.

**Figure 3.**
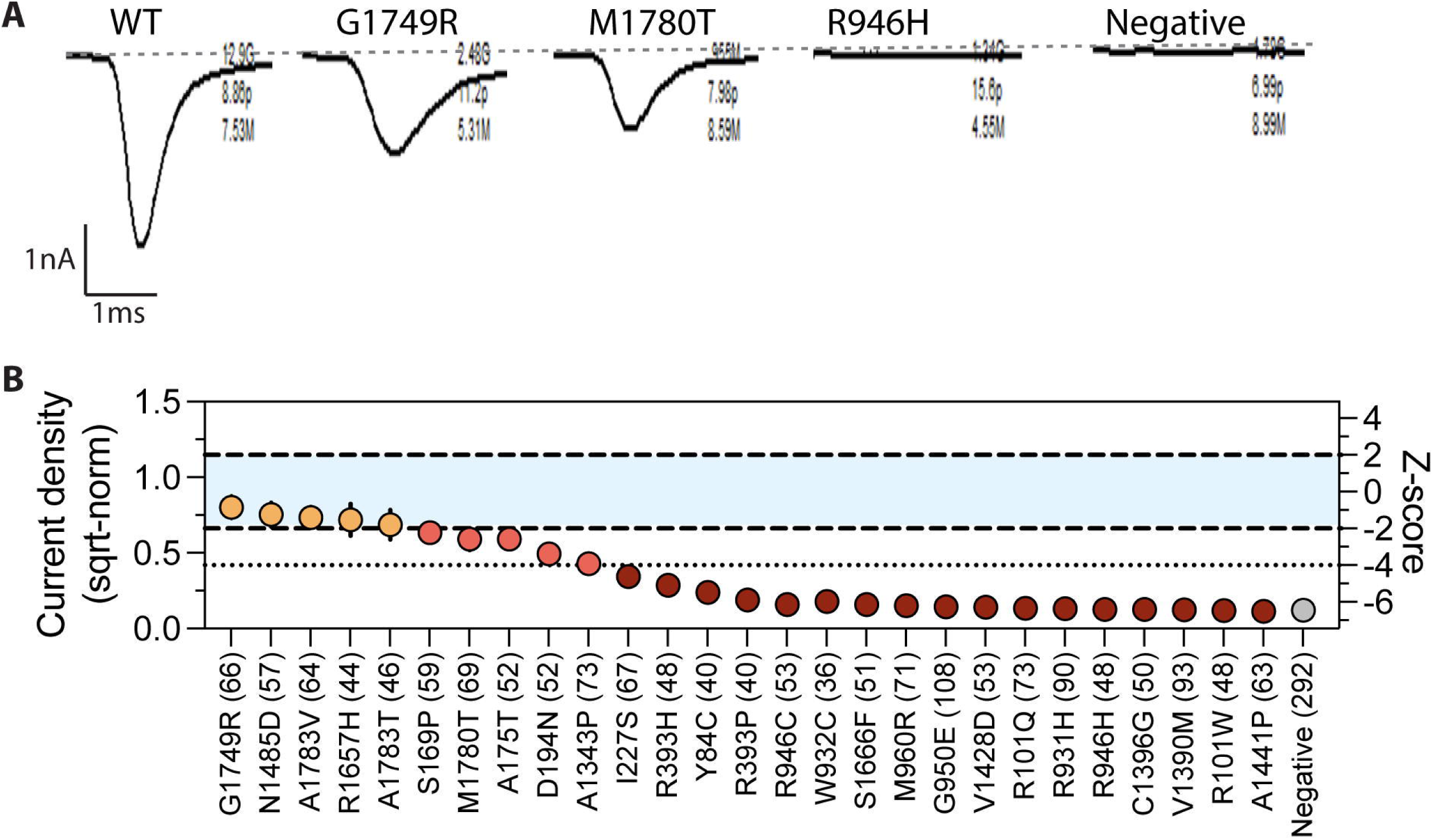
Sensitivity of the *SCN1A* APC assay by current density measurement. **(A)** Representative current traces for selected variants recorded at zero mV. **(B)** Summary of the current density for true pathogenic variants. The normal range is indicated by the blue shade defined by Z = ±2, with the data presented as mean ± 95% CI. Severe LOF variants (Z < –4) are shown in a dark red. Moderate LOF variants (–2 > Z > –4) are shown in red. Variants with current density within the normal range are displayed in orange.

### Voltage-dependence of channel activation and inactivation of Nav1.1

As abnormal channel gating can also contribute to sodium channel dysfunction, the voltage dependence of both channel activation and inactivation were investigated. Example current traces of WT Na_V_1.1 corresponding to channel activation and inactivation are shown in Fig. 4A and C. For the 25 true benign variants, the population mean and standard deviation for channel activation *V*_0.5a_ is –25.9 ± 1.9 mV (Fig. 4D), which was used to define the normal range based on a Z-score of ±2. For the five true pathogenic variants with current density residing within the normal range, two, p.Gly1749Arg and p.Ala1783Val, exhibited a depolarising shift in *V*_0.5a_ (Z = 2.1 and 4.0, respectively, Fig. 4D). The population mean for channel inactivation, *V*_0.5i,_ for WT and the 25 true benign variants was –51.0 ± 1.7 mV (Fig. 4E). For the five true pathogenic variants with current density residing within the normal range, p.Asn1485Asp, p.Arg1657His and p.Ala1783Thr exhibited a hyperpolarising shift in *V*_0.5i_ (Z = –5.1, –4.4 and –6.9, respectively, Fig. 4E). To consider shifts in both *V*_0.5a_ and *V*_0.5i_ as a single parameter, we calculated the *V*_0.5diff_ from the *V*_0.5a_ and *V*_0.5i_ obtained from the same cell for each variant (Fig. 4C). The mean *V*_0.5diff_ for WT and the 25 true benign variants was 25.5 ± 0.8 mV (Fig. 4F). All five pathogenic variants that had current density within the normal range had abnormal *V*_0.5diff_ values (Z_G1749R_ = 4.1, Z_N1485D_ = 14.4, Z_R1657H_ = 8.6, Z_A1783V_ = 12.6, Z_A1783T_ = 8.1, Fig. 4F; orange circles). The remaining five pathogenic variants that showed partial reductions in current density (Fig 4F; red circles), two also have an increased *V*_0.5diff_ (p.Met1780Thr and p.Asp194Asn).

**Figure 4.**
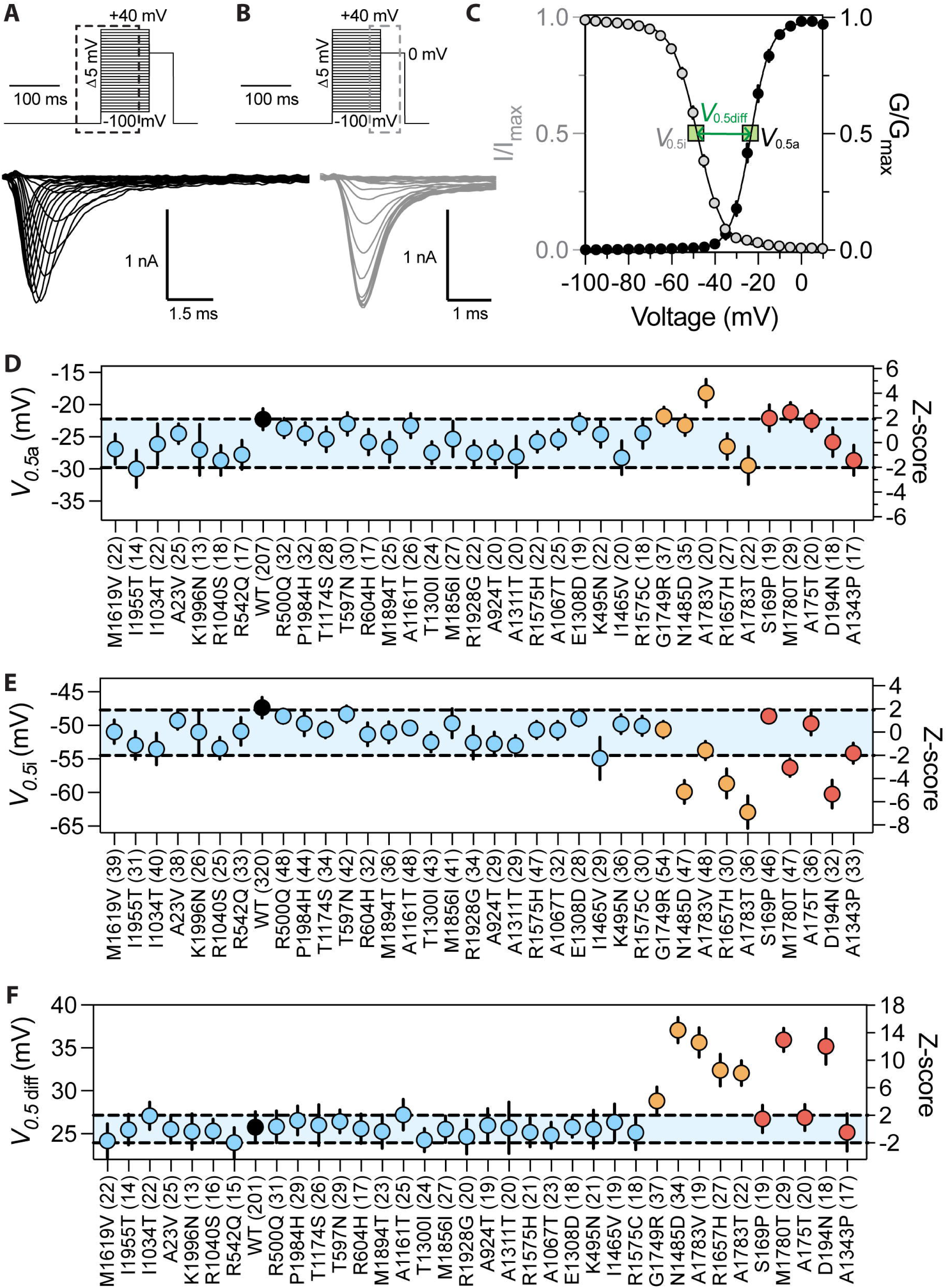
Establishment of parameter *V*_0.5diff_ that encompasses changes in both activation and inactivation. Voltage protocol and representative current traces for **(A)** channel activation highlighted by the black box and **(B)** channel inactivation highlighted by the grey box. **(C)** Normalised WT current (*I*, grey) and WT conductance (*G*, black) plotted against voltage. The voltage corresponding to the midpoint of channel activation (*V*_0.5a_) and inactivation (*V*_0.5i_) is indicated by green squares. The difference between them, *V*_0.5a_ - *V*_0.5i_, is defined as the *V*_0.5diff_. **(D)** The summary of the midpoint of channel activation *V*_0.5a_. **(E)** The summary of the midpoint of channel inactivation *V*_0.5i_. **(F)** The summary of *V*_0.5diff_. In all panels, blue circles are true benign variants, orange circles are true pathogenic variants whose current density falls within the normal range and red circles are true pathogenic variants with moderate reductions in current density (coloured as per Fig. 3). The normal range is indicated by the blue shade defined by Z ±2 and the data is presented as mean ± 95% CI.

### Applying validated *SCN1A* patch-clamp assay to inform variant classification

There were 16 unique *SCN1A* variants identified across 16 families. Forty-eight individuals had seizures. Thirty-two had a GEFS+ phenotype, including; twelve with FS, three with FS and focal seizures, one with FS with combined focal and generalised seizures, two with FS+, three with FS+ and focal seizures, two with FS+ and generalised seizures, one with FS+ and unclassified seizures, four patients with Dravet syndrome, two with focal epilepsy, one with Childhood Absence Epilepsy (CAE) evolving to Juvenile Myoclonic Epilepsy (JME), and one with generalised tonic-clonic seizures alone (GTCA).^32^ An additional ten patients had unclassified epilepsy, four had a single seizure, one had self-limited epilepsy with centrotemporal spikes (SeLECTS), and one with early infantile DEE (EIDEE)^33^. Two variants arose *de novo* (proband of Family C: p.Ser243Phe; proband of Family D: p.Val250Ile); both probands had very early seizure onset at 10 weeks and 23 hours of life, respectively. In addition, the proband from Family C (III.8) is non-ambulant and exhibits a hyperkinetic movement disorder. Detailed clinical phenotypes are provided in Supplementary Table 3.

Prior to functional assessment, 12/16 variants were classified as VUS, three as likely pathogenic and one as likely benign using the ACMG/AMP framework (Supplementary Table 2). In the calibrated *SCN1A* APC assay, five variants show reduced current density whilst 11 had current densities within the normal range (Fig. 5A). Eight of 11 variants with normal range current density had abnormal *V*_0.5diff_ (Fig. 5B). Of the remaining three variants, p.Arg28His is in gnomAD v4.1 with an allele frequency of 5.6E-06 (Fig. 5C and D) and remains classified as likely benign, while p.Cys136Ser and p.Val250Ile are absent from gnomAD v4.1 and retain their pre-assay classifications of VUS and likely pathogenic, respectively. Notably, p.Val250Ile has been reported as pathogenic,^34^ and different missense variants at the same amino acid position in *SCN1A* and other sodium channel genes have been reported as pathogenic and likely pathogenic in ClinVar.

**Figure 5.**
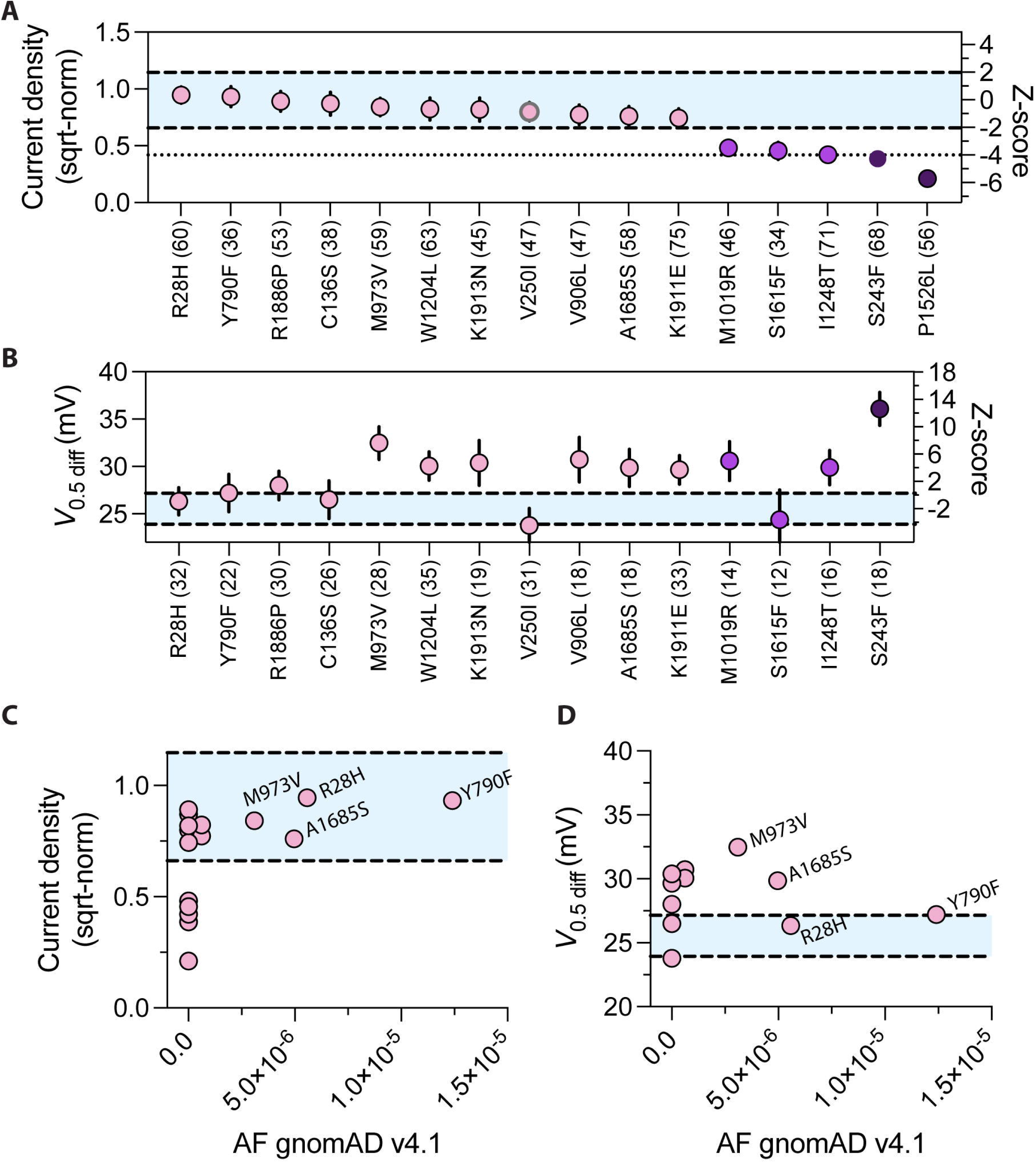
*SCN1A* variants identified in a cohort of epilepsy and febrile seizures from the Epilepsy Research Centre, Melbourne. **(A)** Summary of normalised current density for 16 *SCN1A* variants referred for patch clamp assessment. The darkest shade signifies severe LOF in current density (Z < –4). **(B)** The abnormal gating of variants identified through the use of *V*_0.5diff_. Data is presented as mean ± 95% CI. The gnomAD allele frequency compared to current density **(C)** and *V*_0.5diff_ **(D)**. The normal range is indicated by the blue shade defined by Z of ±2.

Current density and *V*_0.5diff_ were combined within a graded Bayesian framework to derive functional evidence based on a log-likelihood ratio reflecting the magnitude of loss of function (Fig. 6 and Supplementary Table 5). For each variant, the joint current-density and *V*_0.5diff_ profile was compared with the distributions of the true pathogenic and true benign calibration variants to calculate an LLR, with increasing positive LLRs indicating progressively greater support for abnormal function. The resulting LLRs were mapped to the corresponding ACMG/AMP functional evidence strengths, allowing variants with more severe functional abnormalities to receive stronger evidence than variants with partial loss of function. Ten of the 16 variants reached the calibrated evidence threshold for abnormal function on the combined functional score (Fig. 6D). The three remaining variants with an abnormal Z-score on a single readout, p.Tyr790Phe, p.Ser1615Phe and p.Arg1886Pro, did not reach the calibrated likelihood ratio threshold once current density and *V*_0.5diff_ were considered jointly, and PS3 was therefore not applied. Applying PS3 to the ten qualifying variants reclassified six of the 12 VUS as likely pathogenic and upgraded two of the three likely pathogenic variants to pathogenic, a reclassification yield of eight of 16 variants (Fig. 7; Supplementary Table 2). Two further VUS, p.Met973Val and p.Ala1685Ser, received strong functional evidence but remain VUS because the other available criteria are insufficient to reach likely pathogenic, and p.Ser1615Phe and p.Arg1886Pro remain VUS without functional evidence; all four will require additional evidence for definitive classification.

**Figure 6.**
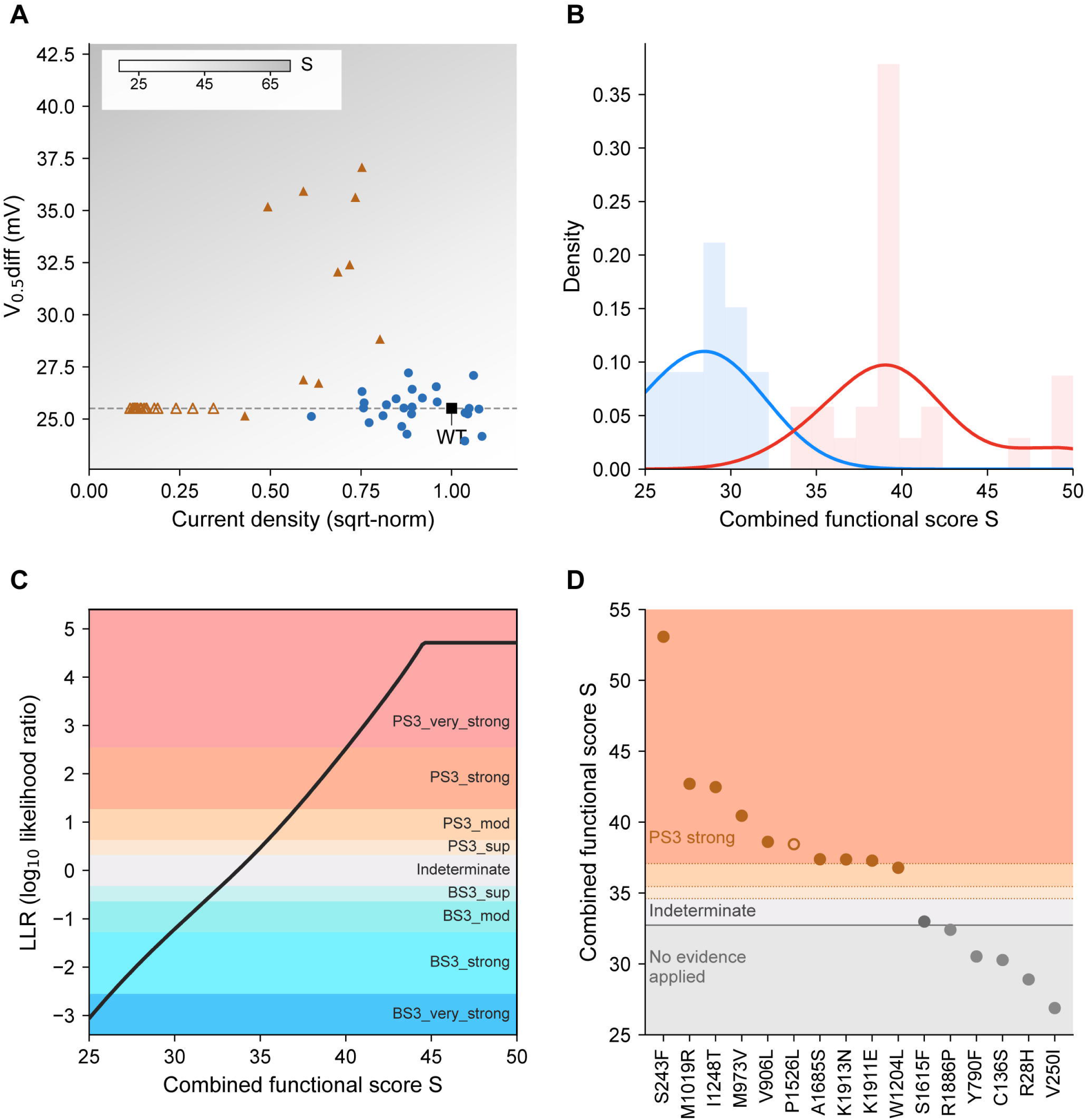
Calibration of the combined functional score and its application to variants of uncertain significance. **(A)** Combined functional score across the two assay readouts. Each point is one variant: true benign variants (blue circles), true pathogenic variants with a measurable window (filled orange triangles), true pathogenic variants for which severe loss of current precluded Boltzmann fitting (open orange triangles, plotted on the dashed line at the WT mean *V*_0.5diff_ of 25.5 mV, the value assigned to them) and WT (black square). The grey field shows the combined functional score S as a function of the two readouts. **(B)** Probability density of S for the benign (blue) and pathogenic (red) reference sets, with the observed scores as histograms. **(C)** Calibrated log-likelihood ratio across the full ClinGen SVI evidence ladder, including very strong. **(D)** Combined functional score for the 16 patient variants, with the evidence actually applied: pathogenic functional evidence (PS3) capped at strong, an indeterminate zone, and a region in which no evidence was applied. Open symbol, variant for which *V*_0.5diff_ was assigned rather than measured.

**Figure 7.**
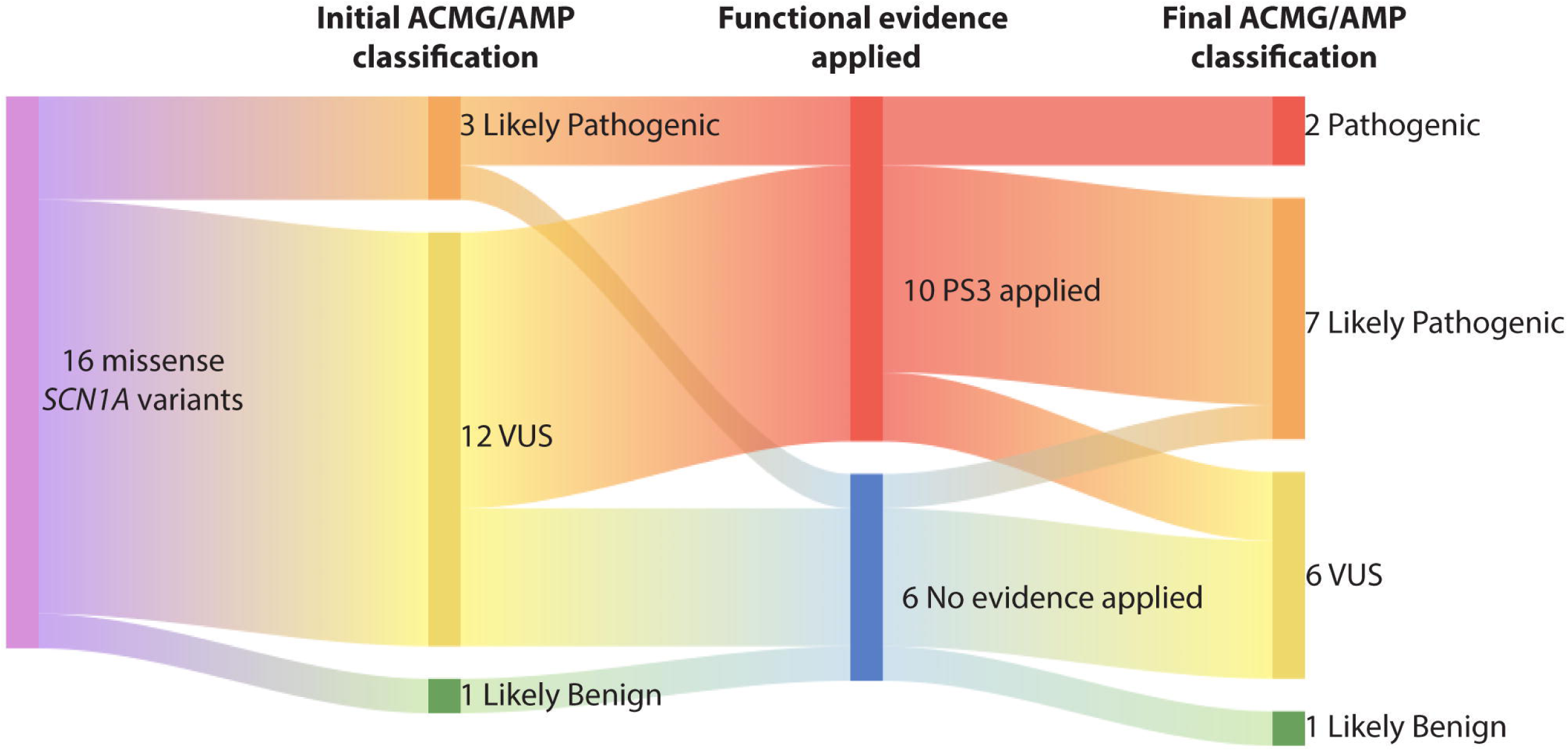
Functional resolution of *SCN1A* variants to determine pathogenicity in individuals with seizure disorders. Sixteen missense *SCN1A* variants detected in patients with epilepsy or febrile seizures were initially classified according to the ACMG/AMP framework supplemented with the latest ClinGen recommendation.^19^ All variants were functionally assessed using our *SCN1A* APC assay. The PS3 criterion was applied to the ten variants whose combined functional score reached the calibrated threshold for abnormal function — at the strong level for nine and at the moderate level for p.Trp1204Leu (see Fig. 6D). PS3 was not applied to the remaining six, which include three variants that were abnormal on a single readout (p.Tyr790Phe, p.Ser1615Phe and p.Arg1886Pro) but did not reach the calibrated threshold once both readouts were considered jointly. The BS3 criterion for normal function was not applied, as recommended by ClinGen.^19^ The criteria used are noted in Supplementary Table 2.

## Discussion

Here, we present a calibrated APC assay that provides functional data for *SCN1A* variants to inform reclassification of VUS based on two parameters: (1) peak current density quantified at zero mV and (2) gating defects using the voltage difference between the midpoints of activation and inactivation (*V*_0.5diff_). These two parameters were combined into a single functional score, calibrated against the true benign and true pathogenic reference variants to yield a log-likelihood ratio and mapped onto the Bayesian evidence-strength thresholds underlying the ACMG/AMP framework,^30,31^ as recommended by the ClinGen SVI Working Group for the graded application of the PS3/BS3 criterion.^18^ The calibrated evidence strength reaches the very strong level for abnormal channel function but was capped at strong for clinical application. This avoids weighting assay-derived pathogenic evidence beyond the level generally supported without additional independent evidence to reach a likely pathogenic classification, whereas normal function evidence (BS3) was not applied.^19^

### Detecting LOF in *SCN1A* variants using *SCN1A* true benign variants

We used a holding potential of –70 mV before measuring the current density at zero mV to mimic the *in vivo* neuronal resting membrane potential.^35^ Furthermore, we considered the function of variants as abnormal when Z < –2 as determined by the mean of all true benign variants, rather than solely relying on the comparison with WT. This ensured that our results were not just statistically significant but also biologically significant.

Reduced current density is a common feature of LOF *SCN1A* variants and is consistent with impaired trafficking of Na_V_1.1 to the cell membrane, with reduced channel availability, or with both; trafficking itself has been assayed directly for only a minority of *SCN1A* variants.^36^ Current density is readily quantified by patch-clamp electrophysiology. Of the 27 SCN1A pathogenic variants associated with Dravet syndrome, 17 had severe LOF (Z < –4), five had moderate LOF (–4 < Z < –2), while five had a current density within the normal range (Z > –2). Gating defects in channel activation or inactivation can also lead to LOF, meaning that abnormal channel function may arise from a trafficking defect, a gating defect or a combination of both.^37,38^ To minimise the parameters necessary for assessing each variant, we merged the changes in *V*_0.5_ for channel activation and inactivation as *V*_0.5diff_. *V*_0.5diff_ with Z > 2 indicate LOF, while *V*_0.5diff_ with Z < –2 may suggest gain-of-function (GOF); however, GOF effects were not specifically examined in this study, since the true pathogenic variants were from patients with Dravet syndrome due to *SCN1A* LOF. The advantage of merging *V*_0.5_ activation and inactivation is that while a slight hyperpolarising shift in activation or depolarising shift in inactivation may not place the variant outside the normal range, the *V*_0.5diff_ allows us to simplify the interpretation of gating parameter changes. There is great utility in the design of such a parameter to distinguish between LOF and GOF. This is particularly useful for genes where information about variant function has serious treatment implications, such as *SCN1A* and the need to avoid sodium channel blockers in LOF disorders. However, further validation for GOF pathogenic variants is required to measure GOF-specific parameters, such as persistent sodium current.

### Dravet syndrome *SCN1A* variants have severe LOF

Dravet syndrome is typically associated with severe *SCN1A* LOF, although other genes have been implicated in rare individuals with Dravet syndrome.^39^ In our set of 27 true pathogenic *SCN1A* variants from Dravet patients, most showed moderate-to-severe reductions in current density, while a minority fell within the normal current range and were identifiable as LOF only via abnormal *V*_0.5diff_, indicating gating defects. In the 16-family cohort, five variants showed LOF by current density, eight were classified as LOF solely by *V*_0.5diff_ and three did not show LOF phenotypes. Clinically, the non-LOF group had focal epilepsy, childhood absence epilepsy evolving to juvenile myoclonic epilepsy and early infantile developmental and epileptic encephalopathy, whereas the LOF group included all patients with Dravet syndrome and those with febrile seizures. p.Tyr790Phe, the remaining variant with a marginally abnormal *V*_0.5diff_, was detected in an individual with self-limited epilepsy with centrotemporal spikes and did not reach the calibrated threshold for functional evidence.

### *SCN1A* Genotype-phenotype correlation

Our study shows clear LOF for the individuals with Dravet syndrome and febrile seizures, reflecting many previous clinico-physiological studies.^40^ It is reassuring that one VUS, one likely benign and one likely pathogenic (possible GOF) detected in other phenotypes are not clearly LOF. This may be because they are due to yet-to-be-discovered pathogenic variants in other genes or because the appropriate type of *SCN1A* dysfunction has not been tested. The best example in our cohort is a man in his 30s with EIDEE, as his phenotype is far more in keeping with the *SCN1A* GOF phenotype: he had seizure onset on the first day of life (23 hours) with tonic seizures and continues to have frequent drug-resistant seizures.^41,42^ He differs from the usual phenotype seen in GOF *SCN1A* diseases as he has mild intellectual disability and a crouch gait, without a movement disorder.

### Limitations

Although HEK293 cells are a well-established expression system for use in APC assays,^14–17,22,43,44^ the effects of both genetic and non-genetic factors such as incomplete penetrance, epigenetics and environmental influences cannot be determined from these cells. While we use variants of known pathogenicity to determine the sensitivity and specificity of our assay, the single-cell measurements do not allow us to examine variant function as part of a neuronal network. Due to these limitations, caution must be exercised when applying functionally normal readouts from our assay to a clinical setting. As a result, we did not apply normal function evidence (BS3) for variants that do not show a LOF phenotype, as outlined in ClinGen’s criteria for classifying neuronal sodium channel variants.^19^ Finally, this assay is applicable only to LOF phenotypes since the calibration used *SCN1A* variants from patients with Dravet syndrome, and cannot be applied to GOF *SCN1A* variants.

## Conclusion

We have validated an APC assay for *SCN1A* missense variants to detect LOF variants on 25 true benign and 27 true pathogenic variants. The assay was employed to evaluate variant function in patients with epilepsy and febrile seizures. Our functional studies have a 50% yield in variant reclassification, with 6/12 VUS reclassified as likely pathogenic and 2/3 likely pathogenic variants upgraded to pathogenic, translating directly to clinical diagnosis and facilitating targeted therapy. To fully realise the benefits of our APC assay, work should be undertaken in close collaboration with research teams, clinical staff and testing laboratories to ensure patients obtain a definitive genetic diagnosis sooner when they have variants with abnormal function.^13^

## Data availability

All data produced in the present study are available upon reasonable request to the authors.

## Acknowledgements

We thank all the patients and their families for participating in our research program. We also acknowledge support from the Victor Chang Cardiac Research Institute Innovation Centre, funded by the New South Wales Government, Australia. This work was made possible through Victorian State Government Operational Infrastructure Support and Australian Government NHMRC IRIISS.

## Ethics

The study was conducted in accordance with the ethical principles of the Declaration of Helsinki. Ethics approval was granted by the Austin Health Human Research Ethics Committee, Melbourne, Australia (approval number H2007/02961).

